# Inequality in access to dental services in a market-based dental care system. A population study from Norway 1975-2018

**DOI:** 10.1101/2021.08.02.21261259

**Authors:** Nan Jiang, Jostein Grytten, Jonas Minet Kinge

**Affiliations:** Department of Community Dentistry, University of Oslo, Oslo, Norway; Department of Obstetrics and Gynecology, Institute of Clinical Medicine, Akershus University Hospital, Lørenskog, Norway; Norwegian Institute of Public Health, Oslo, Norway

## Abstract

**Objective:** To examine income-related inequalities in access to dental services from 1975 to 2018. In Norway, dental care services for adults are privately financed. This might lead to income-related inequalities in access to dental services. However, over the last decades Norwegians have experienced a rapid growth in income, including people with lower income. This may have led to improved access to dental services for these people. Therefore, inequalities in access to dental services may have become less over the last decades.

**Research design:** This was a prospective study. Statistics Norway collected samples of cross-sectional health survey data for the following years: 1975, 1985, 1995, 2002, 2008, 2012 and 2018. For each sample, individuals 20 years and older were drawn randomly from the non-institutionalized adult population using a two-stage stratified cluster sample technique. Inequalities were measured using the concentration index. The dependent variable was use of dental services during the last year and the key independent variable was equalized household income.

**Results:** The concentration index for inequalities in use of dental services according to income decreased from 0.10 (95% confidence interval = 0.09, 0.11) in 1975 to 0.04 (95% confidence interval = 0.03, 0.05) in 2018. The decrease was particularly large from 2002 to 2012. This was a period with a large growth in gross national income.

**Conclusion:** People with a low income had a marked increase in their purchasing power from 1975 to 2018. This coincided with an increase in demand for dental care for this low-income group.

## INTRODUCTION

Equality in access to health services, dental services included, is an important part of Norwegian welfare policy.^1,2^ This policy goal is an important justification for free dental care for children up to the age of 18.^1^ It is undesirable that children’s access to dental services should be limited by their parents’ financial situation. The regular, out-reaching service provided by the public dental service helps to ensure that all children and young people have equal access to dental services, and that inequalities in dental health are minimized.^3^

In Norway, there is no national policy that dental services should be free of charge for adults.^2^ This is different from other Scandinavian countries, where a large part of the costs are subsidized.^4^ In Norway, there has been an ongoing debate since the late 1970s whether dental care for adults should by subsidized by the State.^5-7^ So far, no universal public insurance scheme for dental care has been introduced.

The argument put forward in favour of a subsidy scheme is that it could help to reduce or eliminate inequalities in access to dental services. On the other hand, differences in income in the Norwegian population are relatively small, and among the lowest in the OECD countries.^8^ Further, gross national income (GNI) increased from the early 1970s as a result of the growth of the oil and gas industry.^9,10^ This benefitted everyone, including people with a low income, who were then more able to afford dental services.^8^ This may have led to a reduction in inequalities in access over time, even without a specific dental subsidy scheme.

In the present study, we examined inequalities in access to dental services according to household income over a period of more than 40 years. We analyzed large samples of survey data that were representative of the non-institutionalized adult Norwegian population. The first sample was from 1975, the last sample from 2018. This covers a period in which all people, including people with a low income, had a marked increase in their purchasing power.

## MATERIALS AND METHODS

### The study population

Statistics Norway collected samples of cross-sectional health survey data for the following years: 1975, 1985, 1995, 2002, 2008, 2012 and 2018. For each sample, individuals were drawn randomly from the non-institutionalized population using a two-stage stratified cluster sample technique. The sample sizes and drop-out rates according to year are given in Supplementary Material 1. Statistics Norway has published figures on the representativeness of the samples with respect to gender, age and place of residence (for detailed references to each of the surveys see notes in Supplementary Material 1). For all samples, there were only minor deviations between the sample and the population. These deviations were taken into account in our analyses, using the sample weights given by Statistics Norway. The distribution of the population according to gender, age and place of residence was used to construct the sample weights.

We modified the samples by excluding individuals aged 19 years and younger. Thus, our samples included only individuals who met all the costs for dental services themselves. The sample sizes for the modified samples are given in Supplementary Material 1.

### Variables

The data were collected by personal interviews, using a precoded questionnaire. In all surveys, the respondents were asked when they had last visited the dentist. Several reply options were available, which were slightly different in each survey. For the purpose of the present study, respondents were classified into two groups: those who had visited the dentist during the last year, and those who had last visited the dentist more than a year ago.

Our key independent variable was household income before tax, which consisted of labour income, capital income and all transfers from the Government. All persons who live in Norway have a unique personal identification number. This made it possible to merge the data from the health surveys with the tax records from The Tax Administration of Norway. In our analyses, household income was equivalized to account for household composition using the square root scale.^11^

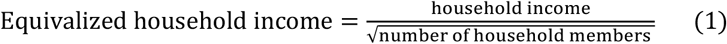

In each survey, some of the respondents had missing information on the variable measuring when they last visited the dentist, and on household income. These respondents were excluded from the analyses. The sizes of the final samples that we analysed, are given in Supplementary Material 1.

### Analyses

To investigate the relationship between use of dental services and household income, we ran the following equation using a linear probability model:

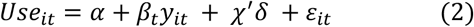

where *Use* is a binary dependent variable taking the value one if an individual, denoted by the subscript *i*, had visited the dentist during the last year, and zero otherwise. *y* is equivalized household income in the calendar year, denoted by subscript *t. χ*^*′*^ is a row vector of age, age squared and gender. We stratified the analysis by calendar year denoted by *t*. To simplify the interpretation of the magnitude of the association between equivalized household income and use of services, we also estimated the following regression:

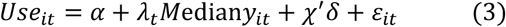

where *Mediany*_*it*_ is a dummy variable that equals one if the individual’s equivalized household income was above the median equivalized household income in the sample, and 0 otherwise.

We measured inequalities using the concentration index, which is commonly used to measure socioeconomic inequality in health and health care utilization.^12,13^ The index is defined with reference to the concentration curve. This curve models the cumulative proportion of individuals who had used dental services against the cumulative proportion of individuals with different levels of household income, ranked from the lowest to the highest. If everyone, irrespective of household income, had exactly the same level of use of services, the concentration curve would be a 45-degree line. This is the line of equality. If level of use of services was higher among those with a high household income, the curve would be below the line of equality, and the concentration index would be positive. The index is in the range -1 to 1. The further the concentration curve is below the line of equality, the closer the index will be to 1.^13^ We calculated the concentration index using the following formula:^14^

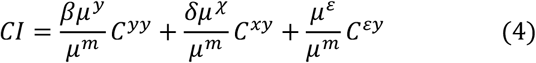

where β and *δ* are the coefficients from Equation (2), *μ*^*y*^ is the mean of the income variable, *μ*^*χ*^ is the mean of the covariates, *μ*^*ε*^ is the mean of the error term, and *μ*^*m*^ is the proportion who had used dental services. *C*^*yy*^, *C*^*xy*^ and *C*^*εy*^ are concentration indices for income, the covariates and the error term, respectively.

The concentration index in Equation (4) takes into account both the direct effect that household income has on use of dental services, and the indirect effects that are transmitted through age and gender.^14^ In our model, the indirect effects will be the component of the association between use and income that is due to the intervening variables age and gender.

For age, there are two negative associations: the association between age and income^15^ and the association between age and use of service.^16^ The indirect effect is the product of these two negative associations. The product of two negative associations is positive. Thus, the indirect effect of income that is transmitted thorough age is positive. For gender, there is one positive and one negative association. The association between gender (female) and use is positive.^17^ The association between gender (female) and income is negative.^18^ The product of one positive and one negative associations is negative. Thus, the indirect effect of income that is transmitted thorough gender is negative.

In the literature, it is common to estimate the partial concentration index.^12,14^ This is a measure of income-related inequality in health after removing the indirect effects of income that are transmitted through age and gender. In our case, the partial concentration index would be a measure of the direct effect of income on use of dental services. We used the method of indirect standardization to estimate the partial concentration index. The estimation was done in three main steps.^14^

First, in one regression we regressed use of dental services on age, and then in another regression we regressed use of dental services on gender. In both regressions, household income was excluded. The regression coefficients from these two regressions included some of the effects of income that were transmitted through age and gender. Second, using the coefficients from these regressions, we computed two concentration indices. One explained inequalities in use of dental services according to age, the other explained inequalities according to gender. Third, we subtracted the two indices from the unstandardized concentration index given in Equation (4). If the unstandardized concentration index is similar to the partial concentration index, then inequalities in use of dental services are a result of a direct effect of income on use. If the indices are different, some of the effects are transmitted thorough age and gender.

The unstandardized and the partial concentration indices measure relative inequality. An alternative approach is to measure absolute inequality, which quantifies the absolute differences in use of dental services between income groups. Wagstaff (2005) and Erregyers (2009) have developed indices that measure absolute inequality.^19,20^ These indices are particularly useful when the outcome is binary, as in our case. With binary outcomes, the minimum and maximum values of the concentration indices depend on the mean of the outcome variable.^19^ This complicates the comparison of the values of the concentration indices across populations in which the mean of the outcome variable varies. In our study, the proportion of individuals who visited the dentist during the last year increased from 1975-2018. Therefore, to take account of the fact that the mean of the outcome variable varied across the samples, we also measured inequalities using the corrected concentration index proposed by Erregyers (2009):

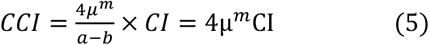

In Equation (5), *μ*^*m*^ is the mean of the outcome variable, *a* and *b* are the maximum and minimum values of the outcome variable, which are 1 and 0 in our case. Hence, the corrected standardized CCI can be written as 4*μ*^*m*^CI.

## RESULTS

### Descriptive statistics

During the period from 1975 to 2018, the percentage of individuals who had visited the dentist during the last year increased from 59.6% to 80.4% (Table 1). During the same time period, the equivalized household income increased from USD 14 551 in 1975 to USD 47 599 in 2018. From 1975 to 2018, the mean number of individuals in the household decreased from 3.1 to 2.4. The mean age of the respondents increased slightly throughout our study period.

**TABLE 1.** Descriptive statistics of the key variables according to year of survey

| Year of survey | Use of dental services during the last year (%) | Number of individuals in the household (median) | Equivalized household income (USD) (median) | Men (%) | Age in years (mean) |
| --- | --- | --- | --- | --- | --- |
| 1975 | 59.6 | 3.1 | 14 551 | 48.4 | 47.2 |
| 1985 | 67.8 | 2.9 | 17 036 | 52.2 | 46.9 |
| 1995 | 66.9 | 2.7 | 26 158 | 51.8 | 49.0 |
| 2002 | 71.7 | 2.6 | 36 645 | 49.8 | 47.4 |
| 2008 | 76.0 | 2.6 | 44 370 | 49.4 | 48.7 |
| 2012 | 79.2 | 2.6 | 47 160 | 52.9 | 48.0 |
| 2018 | 80.4 | 2.4 | 47 599 | 52.2 | 50.9 |

The pattern of use varied according to the respondents’ age, gender and year of the survey (Supplementary Material 2). For those aged 39 or younger, the percentage of individuals who had visited the dentist during the last year decreased from 76.8% in 1975 to 68.4% in 2018. For those 60 or older, the percentage who had visited the dentist increased from 32.9% in 1975 to 88% in 2018. In the age group 40-59 years, the percentage of individuals who had used dental services increased, from 61.0% in 1975 to 82.7% in 2018. For all years, the proportion of men who had visited the dentist during the last year was slightly lower than for women.

### The relationship between use of dental services and household income

For all the years of the survey, equivalized household income was positively associated with use of dental services (Supplementary Material 3). This shows that individuals with an income above the median income in the sample, used more services than those with an income below the median income in the sample. The regression coefficients decreased from 0.14 in 1975 to 0.06 in 2018. The confidence intervals for the later years did not overlap with those for the earlier years. This implies that the association between equivalized household income and use of dental services decreased over time.

### Income-related inequalities in use of dental services over time

Inequalities in use of dental services according to income decreased over time (Table 2). This was a consistent finding, independent of the way in which inequalities were measured.

**TABLE 2.** Different types of concentration indices according to year of survey. 95% confidence intervals in brackets

| Year of survey | Unstandardized<br>concentration<br>index | Partial<br>concentration<br>index | Corrected<br>concentration index<br>(Erregyers) |
| --- | --- | --- | --- |
| 1975 | 0.103<br>[0.092 - 0.114] | 0.088<br>[0.078 - 0.098] | 0.246<br>[0.220 - 0.271] |
| 1985 | 0.093<br>[0.084 - 0.102] | 0.069<br>[0.060 - 0.077] | 0.252<br>[0.228 - 0.276] |
| 1995 | 0.095<br>[0.086 - 0.105] | 0.091<br>[0.081 - 0.101] | 0.257<br>[0.234 - 0.281] |
| 2002 | 0.081<br>[0.068 - 0.094] | 0.082<br>[0.069 - 0.096] | 0.227<br>[0.191 - 0.263] |
| 2008 | 0.053<br>[0.044 - 0.062] | 0.058<br>[0.049 - 0.067] | 0.159<br>[0.134 - 0.184] |
| 2012 | 0.042<br>[0.034 - 0.051] | 0.043<br>[0.035 - 0.052] | 0.134<br>[0.109 - 0.158] |
| 2018 | 0.041<br>[0.033 - 0.050] | 0.038<br>[0.029 - 0.047] | 0.130<br>[0.105 - 0.156] |

During the period 1975 to 2018, the unstandardized concentration index decreased from 0.10 to 0.04. The decrease was particularly large from 2002 to 2008. For the earlier years of the survey, the indices were in the range 0.10 (1975) to 0.08 (2002). For the later years, the indices were in the range 0.05 (2008) to 0.04 (2018). The confidence intervals for the earlier years did not overlap with those for the later years. Over time, the partial concentration index decreased in the same way as the unstandardized concentration index. From 1995 and onwards, the sizes of the two indices were nearly identical. For 1975 and 1985, the partial index was slightly lower than the unstandardized index. For every year of the survey, the confidence intervals for the two indices overlapped. These results show that equivalized household income has a direct effect on use of dental services, but does not have an indirect effect transmitted through age and gender.

For all years, the values for the corrected concentration indices were higher than the values for the unstandardized concentration indices and the partial concentration indices (Table 2). However, during the period 1975 to 2018, the corrected indices decreased in the same way as the other two indices. For example, the decrease was largest at the end of the period. The confidence intervals for the corrected indices for the earlier years did not overlap with those for the later years.

### Income-related inequalities in use of dental services according to age and gender

In Supplementary Material 4, we show the unstandardized concentration indices according to the respondents’ age and gender for the period 1975 to 2018. Below, we first describe the results according to age, then according to gender.

For the age group 20 to 39 years, the index was small. The value was 0.04 for three years (1975, 1994 and 2018). In 1985, the value was 0.02. For the age group 40-59 years, the index decreased from 0.10 in 1975 to 0.02 in 2018. For the age group 60 years or older, there was a marked decrease in the index from 0.19 in 1975 to 0.03 in 2018. The largest decrease for this age group was from 0.13 in 2002 to 0.06 in 2008.

There was no difference in the unstandardized concentration indices according to gender from 1975 to 2012 (Supplementary Material 4). In 2018, the index was 0.06 for men and 0.02 for women. For this year, the confidence intervals for men and women did not overlap.

### Gross National Income and the distribution of income – national figures

There has been a large increase in gross national income (GNI) from 1975 to 2018.^9,10^ In 1975, GNI per capita was NOK 220 400 (deflated by 2015 figures). The corresponding figure in 2018 was NOK 634 800 (Figure 1).

**FIGURE 1.**
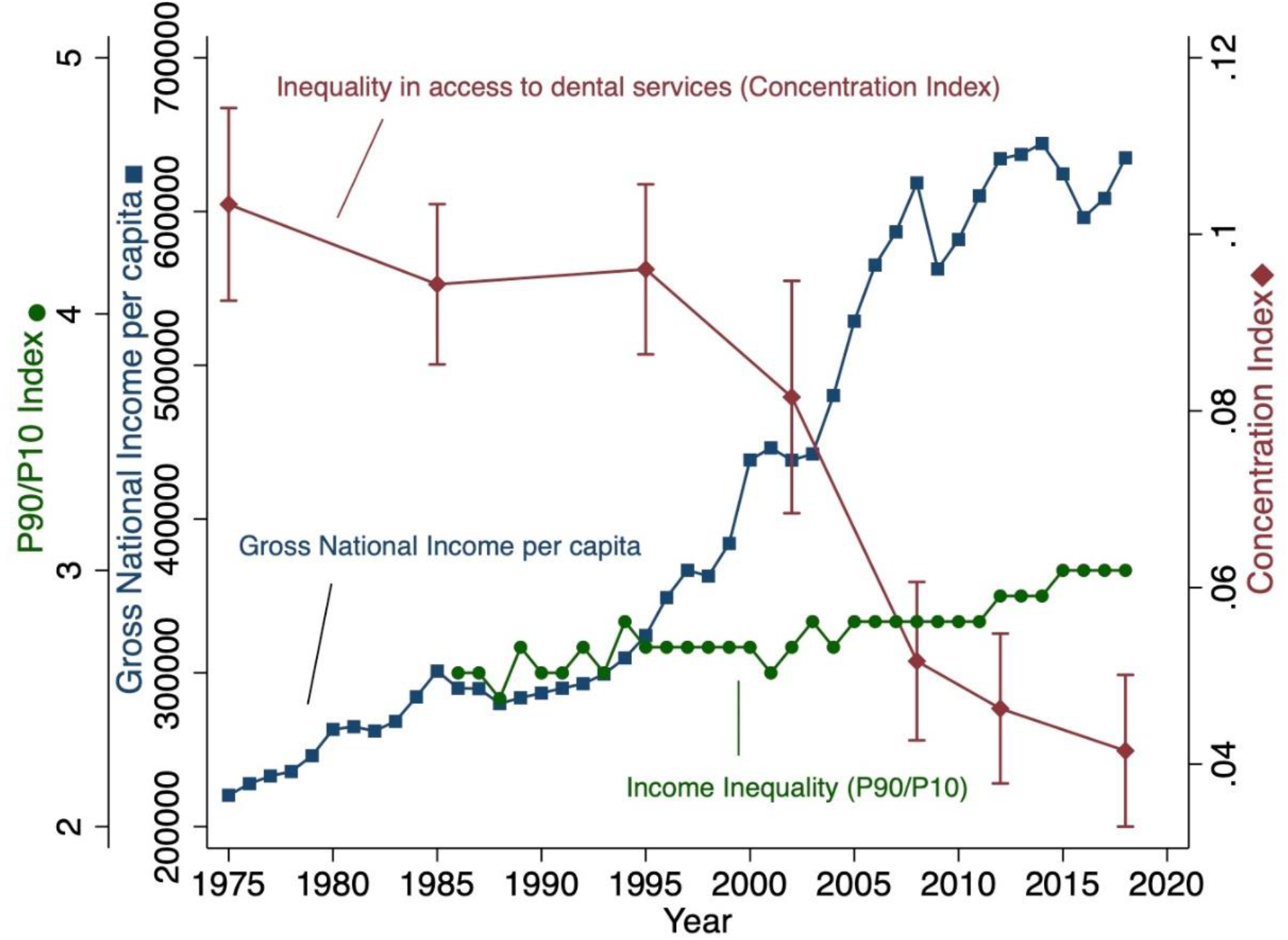
Gross national income per capita in NOK (GNI), income inequality measured by the P90/P10 Index and inequalities in access to dental services measured by the unstandardized concentration index with a 95% confidence interval, according to year.

During the period 1986-2018, the income distribution, measured by the P90/P10 Index showed a fairly stable trend. P90/P10 gives the ratio of the upper value of the ninth decile (= the 10% of people with highest income) to that of the first decile (= the 10 % of people with lowest income).^21^ The exception was the period 2000-2006 in which the P90/P10 increased. This was due to a tax reform, which led to increased inequality due to tax avoiding behaviour among the rich.^22^ For the periods 1986-1999, and 2007-2019 the P90/P10 varied in the range 2.5 (1988) to 3.0 (2015-2018). No official data are available for the P90/P10 Index before 1986.

In Figure 1, we also present the figures for the unstandardized concentration indices for all the years of the survey. This shows the negative relationship between GNI and the concentration indices.

## DISCUSSION

To our knowledge, this is the first study in which inequalities in use of dental services according to income have been examined over a time span of more than 40 years. The analyses were carried out on seven large samples of survey data that were collected in different years. All samples were representative of the non-institutionalized adult population in Norway. This made it possible to describe changes in inequalities in use of dental services at a national level. Previous studies within this field have mainly been carried out at one point in time, or on selected groups of the population, for example in samples of young adults or elderly people.^23-32^

The main finding from this study is that inequalities in use of dental services according to income have decreased over time. This is an interesting result, considering that there is no public or private insurance for dental services in Norway.^2^ Our results can be explained in the following ways:

First, during our study period, there has been a large increase in gross national income.^9,10^ This increase has benefitted all income groups.^8^ Differences in income in the Norwegian population have been relatively small during our whole study period (Figure 1). The increase in gross national income has increased the purchasing power of the population, which has further increased demand for dental care.^33^ Several studies have shown that demand for dental care is responsive to income (for a review see reference number 34). Our results indicate that this response may have been larger for low income groups than for high income groups.

Second, the supply of dentists has been adequate to meet the increase in demand for dental services. The number of dentists in relation to the population is higher in Norway than in most countries in the world.^35,36^ Throughout our whole study period, the number of inhabitants per man-labour year has been about 1100.^37,38^ Dentists are distributed evenly according to region.^39^ The waiting time for a dental appointment is short, less than a week for non-emergency appointments.^40,41^ The dental care market is competitive and most patients can afford dental care at the present level of fees.^42-44^ Thus, when demand for dental services increased for people with a low income, these services were easily available for them.

The concentration index has become a standard measurement tool in studies on equity and inequalities in health care. One limitation of these indices is that they do not have an intuitive interpretation.^12,45^ For example, it is not clear whether an estimated index value reflects a large or a small inequality. This is because the index value is not expressed in natural units. Further, it is not clear whether our index value of 0.10 (1975) is 2.5 times as unequal as 0.04 (2018). This is problematic, since from a dental health policy point of view, we want to know how much dental care should be transferred from the rich to the poor in order to remove all income-related inequalities in use of dental services.

Koolman and van Doorslaer (2004) have suggested that the index value can be given a meaningful interpretation by multiplying it by 0.75.^46^ In our case, this gives the percentage of users of dental services that would need to be redistributed from the richer half of the population to the poorer half of the population in order to arrive at a distribution with an index value of zero, i.e. no inequality.^12^ For 2018, 3% of users would have to be redistributed. This is a small percentage, which indicates that inequalities in access to dental services are not a serious problem, even without a subsidy scheme.

Income-related inequalities in use of dental services for the age group 20-39 years were particularly small (Supplementary Material 4). This was the case for all the years of the survey. Most likely, this is because free dental care in childhood has contributed to regular use of dental services in adulthood. In Norway, well over 90 per cent of children and young adults under 19 years of age have annual appointments with a public dentist.^38^ All their dental treatment is free, and they receive information and guidance about how to prevent dental disease.^4^ This means that positive dental behaviour can be established in childhood and lead to good oral health in adult life.^47,48^ This has contributed to an increase in regular use of dental services for all people in the age group 20-39 years, independent of their level of income.

In conclusion, we found that from the beginning of our study period in 1975 until the end in 2018, differences in use of dental services according to income decreased. The study was carried out in a population for which there was no public or private insurance for dental treatment. All dental treatment had to be paid for by the patient. A possible explanation for our finding is that people with a low income had a marked increase in their purchasing power from the early 1970s and onwards. This resulted in increased demand for dental care.

### SM

Supplementary Material 1.docx

## Data Availability

Data are available from Norwegian Centre for Research Data

https://www.nsd.no/en

## Acknowledgements

The project was approved by the Norwegian Centre for Research Data (order number: 60). We wish to thank Linda Grytten for language correction and Irene Skau for technical assistance.

