## Supplementary file for "Inequality in access to dental services in a market-based dental care system. A population study from Norway 1975-2018"

**TABLE S1** Sample sizes according to year

|  | All ages | | | | | |  | 20 years and older | | | | | |
| --- | --- | --- | --- | --- | --- | --- | --- | --- | --- | --- | --- | --- | --- |
|  | Gross sample^1^ |  | Non-response | |  | Net sample^2^ |  |  |  | Missing information on key variables^3^ | |  |  |
| Year of survey | Number of respondents |  | Number of respondents | Per cent |  | Number of respondents |  | Number of respondents |  | Number of respondents | Per cent |  | Number of respondents used in the analyses |
| 1975 | 12 438 |  | 1 424 | 11.5 |  | 11 014 |  | 7 510 |  | 311 | 4.1 |  | 7 199 |
| 1985 | 13 438 |  | 2 862 | 21.3 |  | 10 576 |  | 7 444 |  | 211 | 2.8 |  | 7 233 |
| 1995 | 14 000 |  | 3 752 | 26.8 |  | 10 248 |  | 7 474 |  | 2 | 0.0 |  | 7 472 |
| 2002 | 4 839 |  | 1 442 | 29.8 |  | 3 397 |  | 3 195 |  | 1 | 0.0 |  | 3 194 |
| 2008 | 9 684 |  | 3 219 | 33.2 |  | 6 465 |  | 6 008 |  | 25 | 0.4 |  | 5 983 |
| 2012 | 11 387 |  | 5 201 | 44.4 |  | 6 186 |  | 5 795 |  | 132 | 2.3 |  | 5 663 |
| 2018 | 11 393 |  | 5 412 | 47.5 |  | 5 981 |  | 5 667 |  | 418 | 7.4 |  | 5 203 |
| Notes. All surveys were carried out by Statistics Norway, Oslo and were made available by NSD - Norwegian centre for research data. | | | | | | | | | | | |  |  |
| The references to the surveys are as follows: | | | |  |  |  |  |  |  |  |  |  |  |
| Health Survey 1975 (https://doi.org/10.18712/NSD-NSD0015-V3) | | | | |  |  |  |  |  |  |  |  |  |
| Health Survey 1985 (https://doi.org/doi:10.18712/NSD-NSD0016-V4) | | | | |  |  |  |  |  |  |  |  |  |
| Health Survey 1995 (https://doi.org/10.18712/NSD-NSD0349-V4) | | | | |  |  |  |  |  |  |  |  |  |
| Level of Living 2002 - Cross sectional study - Health (https://doi.org/10.18712/NSD-NSD0669-V4) | | | | | | | | |  |  |  |  |  |
| Level of Living 2008 - Cross sectional study - Health (https://doi.org/10.18712/NSD-NSD1327-V7) | | | | | | | | |  |  |  |  |  |
| Level of Living EU SILC, 2012 (https://doi.org/10.18712/NSD-NSD1967-V2 ) | | | | | | |  |  |  |  |  |  |  |
| Level of Living EU SILC, 2018 (https://doi.org/10.18712/NSD-NSD2671-V6) | | | | | | |  |  |  |  |  |  |  |
| ^1^ Number of individuals who were asked to participate | | | |  |  |  |  |  |  |  |  |  |  |
| ^2^ Number of individuals who actually participated | | | |  |  |  |  |  |  |  |  |  |  |
| ^3^ Equivalized household income and use of dental services during the last year | | | | | | |  |  |  |  |  |  |  |

**TABLE S2.** Use of dental services according to age and gender for the years 1975, 1995 and 2018

| Independent variables | | 1975 Use of dental services | | |  | 1995 Use of dental services | | |  | 2018 Use of dental services | | |
| --- | --- | --- | --- | --- | --- | --- | --- | --- | --- | --- | --- | --- |
|  |  | Yes (%) | No (%) | Total (n) |  | Yes (%) | No (%) | Total (n) |  | Yes (%) | No (%) | Total (n) |
| Age (in years) | |  |  |  |  |  |  |  |  |  |  |  |
|  | 20-39 | 76.8 | 23.2 | 2 749 |  | 67.9 | 32.1 | 2 664 |  | 68.4 | 31.6 | 1 510 |
|  | 40-59 | 61.0 | 39.0 | 2 543 |  | 78.4 | 21.6 | 2 660 |  | 82.7 | 17.3 | 1 963 |
|  | ≥ 60 | 32.9 | 67.1 | 1 907 |  | 51.4 | 48.6 | 2 148 |  | 88.0 | 12.0 | 1 776 |
| Gender | |  |  |  |  |  |  |  |  |  |  |  |
|  | Men | 56.7 | 43.3 | 3 481 |  | 65.3 | 34.7 | 3 599 |  | 78.6 | 21.4 | 2 740 |
|  | Women | 62.2 | 37.8 | 3 718 |  | 68.3 | 31.7 | 3 873 |  | 82.4 | 17.6 | 2 509 |

**TABLE S3.** The relationship between equivalized household income^1^

and use of dental services according to year of survey

| Year of survey | Regression coefficient [95% confidence interval] |
| --- | --- |
| 1975 | 0.145*** |
|  | [0.124 - 0.166] |
| 1985 | 0.104*** |
|  | [0.082 - 0.125] |
| 1995 | 0.117*** |
|  | [0.095 - 0.138] |
| 2002 | 0.120*** |
|  | [0.090 - 0.155] |
| 2008 | 0.087*** |
|  | [0.065 - 0.110] |
| 2012 | 0.085*** |
|  | [0.064 - 0.107] |
| 2018 | 0.067*** |
|  | [0.044 - 0.090] |

^1^ Equals 1 if the individual’s equivalized household income was above the median

equivalized household income in the sample, and 0 otherwise.

*** p<0.001

**TABLE S4.** The unstandardized concentration index according to age, gender and year of survey.

95% confidence intervals in brackets

|  | Age (in years) | | |  | Gender | |
| --- | --- | --- | --- | --- | --- | --- |
| Year of survey | 20 -39 | 40 -59 | ≥ 60 |  | Men | Women |
| 1975 | 0.042 | 0.101 | 0.194 |  | 0.102 | 0.108 |
|  | [0.030 - 0.054] | [0.083 - 0.119] | [0.157 - 0.230] |  | [0.085 - 0.118] | [0.094 - 0.122] |
| 1985 | 0.023 | 0.058 | 0.155 |  | 0.099 | 0.095 |
|  | [0.012 - 0.034] | [0.044 - 0.072] | [0.129 - 0.181] |  | [0.085 - 0.113] | [0.083 - 0.106] |
| 1995 | 0.041 | 0.042 | 0.182 |  | 0.099 | 0.097 |
|  | [0.025 - 0.058] | [0.029 - 0.055] | [0.162 - 0.203] |  | [0.084 - 0.113] | [0.085 - 0.110] |
| 2002 | 0.050 | 0.055 | 0.136 |  | 0.078 | 0.086 |
|  | [0.026 - 0.075] | [0.038 - 0.072] | [0.109 - 0.163] |  | [0.059 - 0.097] | [0.068 - 0.104] |
| 2008 | 0.053 | 0.035 | 0.064 |  | 0.052 | 0.058 |
|  | [0.033 - 0.073] | [0.023 - 0.047] | [0.049 - 0.079] |  | [0.038 - 0.065] | [0.047 - 0.070] |
| 2012 | 0.033 | 0.037 | 0.049 |  | 0.050 | 0.037 |
|  | [0.015 - 0.050] | [0.025 - 0.049] | [0.036 - 0.062] |  | [0.038 - 0.062] | [0.026 - 0.048] |
| 2018 | 0.040 | 0.025 | 0.032 |  | 0.063 | 0.025 |
|  | [0.020 - 0.061] | [0.012 - 0.038] | [0.020 - 0.044] |  | [0.050 - 0.075] | [0.013 - 0.037] |
